# Bayesian Estimation of real-time Epidemic Growth Rates using Gaussian Processes: local dynamics of SARS-CoV-2 in England

**DOI:** 10.1101/2022.01.01.21268131

**Authors:** Laura M Guzmán-Rincón, Edward M Hill, Louise Dyson, Michael J Tildesley, Matt J Keeling

**Affiliations:** The Zeeman Institute for Systems Biology & Infectious Disease Epidemiology Research, School of Life Sciences and Mathematics Institute, University of Warwick, Coventry, CV4 7AL, United Kingdom; Joint UNIversities Pandemic and Epidemiological Research, https://maths.org/juniper/

**Keywords:** Bayesian hierarchical modelling, epidemiological trends, Gaussian processes, growth rate estimation, public health tools, spatial heterogeneity

## Abstract

Quantitative assessments of the recent state of an epidemic and short-term projections into the near future are key public health tools that have substantial policy impacts, helping to determine if existing control measures are sufficient or need to be strengthened. Key to these quantitative assessments is the ability to rapidly and robustly measure the speed with which the epidemic is growing or decaying. Frequently, epidemiological trends are addressed in terms of the (time-varying) reproductive number *R*. Here, we take a more parsimonious approach and calculate the exponential growth rate, *r*, using a Bayesian hierarchical model to fit a Gaussian process to the epidemiological data. We show how the method can be employed when only case data from positive tests are available, and the improvement gained by including the total number of tests as a measure of heterogeneous testing effort. Although the methods are generic, we apply them to SARS-CoV-2 cases and testing in England, making use of the available high-resolution spatio-temporal data to determine long-term patterns of national growth, highlight regional growth and spatial heterogeneity.

## 1 INTRODUCTION

Statistical analysis of the SARS-CoV-2 pandemic has been instrumental in both assessing the current status of infection at a local or national level (The Royal Society, 2020; Hellewell et al., 2020; Flaxman et al., 2020; Davies et al., 2020, 2021), and extrapolating to generate short-term projections. Arguably good statistical knowledge is key to the control of epidemics, as it provides a quantitative assessment of control measures and can highlight sectors of the population in which additional targeted controls may be needed. Five elements combine to make the statistical analysis of the SARS-CoV-2 pandemic difficult: many infections are asymptomatic and go undetected; the regular use of lateral flow devices, which would detect asymptomatic infection, is heterogeneous across time, space and age-groups; the use of polymerase chain reaction (PCR) testing (adopted as the gold-standard in the UK) also changes across time and space, presumably as individuals react to changes in perceived risk; infection and testing are inherently stochastic processes; and there are distributed lags between infection and detection. These five factors mean that the prompt identification of rising infection (especially in relatively small populations) requires sophisticated statistical methods.

The reproductive number, *R*, has gained substantial media and political interest during the SARS-CoV-2 pandemic as a simple statistical indicator of the current epidemiological trends, with *R* < 1 corresponding to a declining outbreak and *R* > 1 corresponding to a growing outbreak (Vegvari et al., 2021). During 2020 in England, the nationwide estimate of *R* (UK Health Security Agency, 2020b) was one of the key metrics in determining the national alert level with implications for changes in control measures (UK Health Security Agency, 2020a); hence placing great political, economic and public-health importance on this single value. A robust and rapid estimation of *R* (or the epidemic growth rate *r*), together with levels of uncertainty, remains a key public-health tool. The estimation needs to be rapid, such that prompt action can be taken before the burden on health services becomes too great; the estimation also needs to be robust, as the economic and social consequences of action can be costly and so should only be enacted when there is considerable certainty that such measures are needed. As such there is a clear need for continued development of statistical methods that can extract a meaningful signal from complex and noisy epidemiological data.

Obtaining an accurate and timely measure of *R* generally requires a robust estimate of either the generation time or the infectiousness profile over time (Wallinga and Lipsitch, 2007) (capturing the expected level of transmission at time *t* after infection). Both of these necessitate detailed individual-level observations (Hart et al., 2021; Abbott et al., 2020, 2021) and may therefore be context dependent, leading to a diversity of *R* estimates from the same population-level data (Funk et al., 2020). Here, we adopt the more parsimonious approach of working with the growth rate *r* (such that the number of infections grows like *I* (*t*) ∼ *exp* (*r t*)), in which case our threshold for a growing or declining outbreak becomes where *r* is greater or less than zero, respectively.

Given the importance of real-time estimation of the growth rate, *r*, or the reproductive number, *R*, multiple statistical methods have been developed (The Royal Society, 2020; Gostic et al., 2020). All methods have advantages and potential problems, with an inevitable trade-off between robustness and timeliness. Most naively, the growth rate can be estimated by simply measuring the rate of change of log(infection), where infections are often approximated as being proportional to reported cases. This naive approach is confounded by the stochastic nature of transmission and reporting, requiring either smoothing of the data or fitting the growth rate over a defined time window - longer windows and more smoothing eliminate stochastic effects, but mean that real-time estimates of the growth rate and *R* are considerably lagged. The UK government dashboard (UK Health Security Agency, 2020c) expands on these simple ideas to produce estimates of the growth rate at the national scale, calculated as the relative change over seven days in the smoothed number of cases (smoothed by taking a mean over a seven-day window). In recent years, EpiEstim (Cori et al., 2013) has grown in popularity as a method of estimating changing *R* values, due to its flexibility and accuracy (Funk et al., 2020). EpiEstim uses a Bayesian framework to compare the reported number of cases over a time window with the projection based on the infectiousness profile and historic reporting to generate an estimate of *R* in a given window.

In this paper we develop a novel method to generate a real-time estimate of the growth rate of infection in small stochastic populations. Our flexible method uses a Bayesian approach to compute the posterior distribution of the growth rate at any point in time and produces samples of the joint posterior distribution of the growth rate for any given interval. We use Gaussian processes (GPs) to fit to the reported data, which gives us flexibility in smoothing the count of new cases according to the GP parameters. We fit to two different measures: the raw number of recorded cases in a region, as defined by PCR positives in the community; or the proportion of community PCR tests that are positive. The latter provides a more stable estimate when testing patterns are changing rapidly.

We first outline the basic methodology and illustrate its use on surrogate data sets where the growth rate is known. We then apply our model to data on SARS-CoV-2 cases in England, initially at a national-level by estimating the daily growth rate of SARS-CoV-2 from 1st September 2020 to 6th December 2021 (as available at the time of writing). Finally, we explore the spatial heterogeneity in cases at the Lower Tier Local Authority (LTLA) level in April 2021 when the Delta variant was increasing in the North-West of England - a time when the spatial variability of epidemic behaviour was key to understanding the impact of the new variant.

## 2 METHODS

We describe a model framework to estimate the growth rate, *r*, of an epidemic based on the count of reported infections (cases). If the counts are recorded through a testing programme, to adjust for changes in testing effort the model can also incorporate the total number of tests performed over time. We assume that the underlying process generating the count of cases is given by a one-dimensional Gaussian process (Section 2.1), and obtain the growth rate by sampling from the derivative of the process (Section 2.2).

### 2.1 Model structure

For a given community, let 𝒯 = {1, …, *T* } be a set of time indices at which data are collected. For each *t* ∈ 𝒯, *y*_*t*_ denotes the number of positive test results at time *t* and *n*_*t*_ denotes the total number of tests. In the context of SARS-CoV-2, data are generally collated daily with the potential for missing data, which our proposed models allow for; for other infections data may be collected over larger or irregularly spaced intervals.

#### 2.1.1 Positives model

We first propose the following Bayesian hierarchical model, labelled as the *positives model*, which only requires knowledge of *y*_*t*_ (the number of positive cases at time *t*) and is therefore applicable in situations where *n*_*t*_ (the number of tests at time *t*) is not available. The model assumes that *y*_*t*_ follows a negative binomial distribution parameterised by its mean *μ*_*t*_ and a time-homogeneous over-dispersion parameter *η*. The probability mass function of *y*_*t*_ under this parameterisation is:

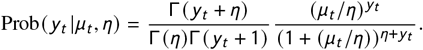

The parameter log(*η*) is assigned a normal prior 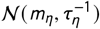. The log relative risk, log(*μ*_*t*_), is decomposed into the sum of a smooth term *x*_*t*_ and a Gaussian error term *ϵ*_*t*_ with zero mean and precision *τ*_*ϵ*_ ∼ Γ(*a*_*ϵ*_, *b*_*ϵ*_). The model can therefore be expressed as:

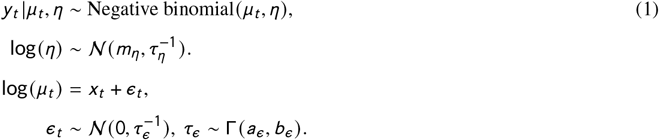

where the hyperparameters underpinning the distribution of the error precision (*a*_*ϵ*_ and *b*_*ϵ*_) and the over-dispersion hyperparameters (*m*_*η*_ and 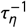) are specific to the problem and quoted in the results. To avoid identifiability issues with the terms *x*_*t*_, we impose a sum-to-zero constraint to the error terms such that Σ_*t*_ *ϵ*_*t*_ = 0.

The prior on the smooth terms *x*_*t*_ is given by a Gaussian process *f* on ℝ such that *x*_*t*_ = *f* (*t*), where *f* has mean zero, covariance *k*_***θ***_ (*f* (*s*), *f* (*s*^′^)) between the value of the process *f* at times *s* and *s*^′^ and hyperparameters ***θ***:

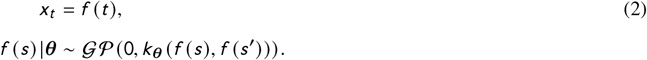

A comprehensive summary of such regression models using Gaussian processes can be found in (Rasmussen and Williams, 2006, Ch. 2). Here, we use a one-dimensional Matérn covariance family (Stein, 1999), since the resulting process *f* is stationary and isotropic, and the smoothness can be specified through a single smoothing parameter *ν*. We choose *ν* = 3/2 which results in the covariance function:

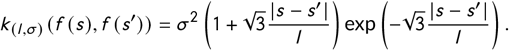

which also depends on the additional hyperparameters ***θ*** = (*l, σ*); where *l* is the length-scale, and *σ*^2^ is the marginal variance of the process. We set the joint prior of *l* and *σ* as (log(*l*), log(*σ*)) ∼ 𝒩 ((*l*_0_, *σ*_0_), *B* ^−1^) where *l*_0_ and *σ*_0_ are baseline values for the length-scale and precision, respectively, and *B* is the precision matrix of the joint prior. A diagram of the model is shown in S.1 of the Supplementary material.

This basic model can be extended by adding additional terms which capture other elements of the dynamics. For example, reported SARS-CoV-2 cases in England have a pronounced day-of-the-week effect, with fewer cases reported on weekends (see Figure 2A). The day-of-the-week effect (which is included in all the results shown below) can be interpreted as an alternative error term with possible non-zero mean whose parameters are allowed to depend on the day of the week. Formally, we substitute the error term *ϵ*_*t*_ by a day-of-the-week effect *w*_*d*(*t*)_ with a Gaussian hyperprior with zero mean and precision *τ*_*w*_ ∼ Γ(*a*_*w*_, *b*_*w*_) (where *d* (*t*) is the day of the week on *t*). We impose a sum-to-zero constraint Σ_*t*_ = *w*_*t*_ = 0 to avoid identifiability issues with the terms *x*_*t*_.

#### 2.1.2 Proportions model

If the number of tests is known, then an alternative model formulation is possible that accounts for changes in testing behaviour over time; we label this model the *proportions model* and seek to capture the proportion of tests that are positive. In this case, *y*_*t*_ (the number of positive cases at time *t*) given *n*_*t*_ (the number of tests at time *t*) is assumed to follow a beta-binomial distribution with mean parameter *μ*_*t*_, over-dispersion parameter *ρ* and number of trials *n*_*t*_. We use a beta-binomial distribution to account for both the bounded nature of *y*_*t*_ (which is bounded above by *n*_*t*_) and the over-dispersion.

The probability mass function of *y*_*t*_ under this parameterisation is given by:

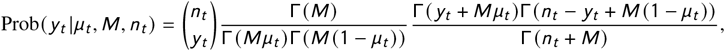

where *M* = (1/*ρ*) − 1. Given the bounded nature of the positive tests, such that *μ*_*t*_ ∈ (0, 1), we utilise the inverse logit transform (logit^−1^), and assume that logit^−1^ (*μ*_*t*_) is decomposed into the sum of a smooth term *x*_*t*_ and a Gaussian error term *ϵ*_*t*_ with zero mean and precision *τ*_*ϵ*_ ∼ Γ(*a*_*ϵ*_, *b*_*ϵ*_). The transformed over-dispersion parameter logit^−1^ (*ρ*) is assigned a normal prior 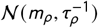. As in the positives model, the prior on *x*_*t*_ is given by the Gaussian process described in Section 2.1.1:

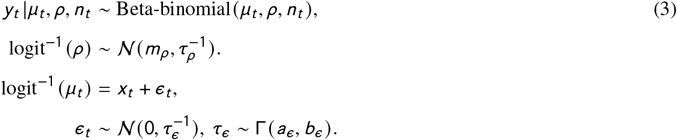

### 2.2 Growth rate sampling

The instantaneous growth rate is defined as the *per capita* change in the number of new cases per time period. That is, if *w*_*t*_ is the process generating new cases at time *t*, the growth rate corresponds to *r*_*t*_ = *∂*_*t*_ (*w*_*t*_)/*w*_*t*_, or equivalently, *r*_*t*_ = *∂*_*t*_ (log *w*_*t*_); where *∂*_*t*_ signifies the time derivative. However, *w*_*t*_ is unknown in practice, so we instead approximate the growth rate using our fitted Gaussian process. For the positives model, we approximate *r*_*t*_ as the growth rate of the process fitting the number of new reported cases, exp(*f* (*t*)). That is, 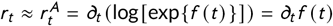 and therefore, *r*_*t*_ can be estimated as the derivative of the Gaussian process *f*. For the proportions model, we approximate *r*_*t*_ as 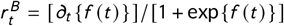, such that 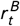 corresponds to the growth rate of new reported cases minus the growth rate the new tests performed (see S.2 of the Supplementary material).

To capture the inherent uncertainty in the process *f*, we sample from the derivative of the process *f* to obtain samples of the growth rate. Note that Gaussian processes with the Matérn covariance are mean-square differentiable if *ν* > 1, which is satisfied by our choice of *ν* = 3/2 (Stein, 1999). We obtain samples of the derivative by taking numerical approximations of the derivative (*∂*_*t*_) of samples drawn from the process *f*. That is, for a given sample *g* of *f*, we approximate the derivative as 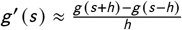 with error 𝒪 (*h*), where *h* is the window size of the approximation.

### 2.3 Implementation

We implement the model in R using the package `INLA` (Rue et al., 2009), where the posterior distribution of the parameters of the model is obtained using a Laplace approximation. The Gaussian process with Matérn kernel is computed as the solution of a stochastic partial differential equation (Lindgren et al., 2011), obtained by the Finite Element Method (FEM). To fit the model using the FEM implementation in `INLA`, we create a one-dimensional mesh with equally-spaced nodes that represent time points. The nodes are located according to the frequency of reported counts; that is, if the data is reported daily, one node per day is located, even if there is missing data. To avoid boundary effects, the mesh domain is extended by at least the length of the studied period (extra nodes are added before the first observation and after the last observation) (Lindgren and Rue, 2015). The code is available in GitHub/juniper-consortium/growth-rate-estim. For the rest of the paper, we use weakly informative priors to the over-dispersion parameters of the models, such that log(*η*) ∼ 𝒩 (0, 0.01^−1^) and logit^−1^ (*ρ*) ∼ N(0, 0.5^−1^). More restrictive priors could reject values of the hyperparameters possibly explained by the data (Rasmussen and Williams, 2006, Ch. 5). Choices of *τ*_*ϵ*_, *l*_0_, *σ*_0_ and *B* are case-specific and are detailed in the results.

### 2.4 Model validation

To validate the accuracy of the models, we generate synthetic epidemiological data from a single homogeneous population of size *N* = 1, 000, 000. We assume for the first 100 days there is an underlying growth rate of *r* = 0.03 per day. For the second 100 days, we assume that controls are enacted and the epidemic goes into decline with a rate of *r* = −0.02. More precisely, the number of infections *y* (*t*) on day *t* are sampled from a Poisson distribution with rate 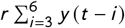 for *t* > 6 and exp(*r t*) for *t* <= 6 (where *r* = 0.03 for *t* <= 100, *r* = −0.02 for *t* > 100, and *y* (0) = 100).

We compare two scenarios for the number of daily tests *n* (*t*). As our purpose is to test the model accuracy under a known growth rate, rather than discuss the effect of the test sampling, we make highly optimistic assumptions for the frequency of testing. In the first scenario, a random ten percent of the population is tested daily, *n* (*t*) = 0.1*N* ; in the second scenario, tests increase linearly from *n* (0) = 0.01*N* to *n* (200) = 0.1*N*.

We run both the positives model and proportions models for each scenario. We set *l*_0_ = 50, *σ*_0_ = 1, and impose *B* = to have non-informative priors for the parameters of the Gaussian process (where is the identity matrix). For the approximation of the derivative, we set a window of *h* = 3 days for all times except near the boundary, where we choose *h* = 1 for *t* = 1, 200, and *h* = 2 for *t* = 2, 199.

For simulation 1, with a constant daily testing rate, for both models the true growth rate is in the posterior credible interval (CI) for all time steps, except near *t* = 100 (Figure 1, top row). The lack of abrupt transition at the *t* = 100 breaking point is due to the smoothness of the Gaussian process (as captured by the assumed length scale, *l*_0_). Although we could use a less smooth covariance function, such a covariance function choice would overfit the data, responding to small stochastic variations and hence not capturing the true underlying growth rate.

**FIGURE 1.**
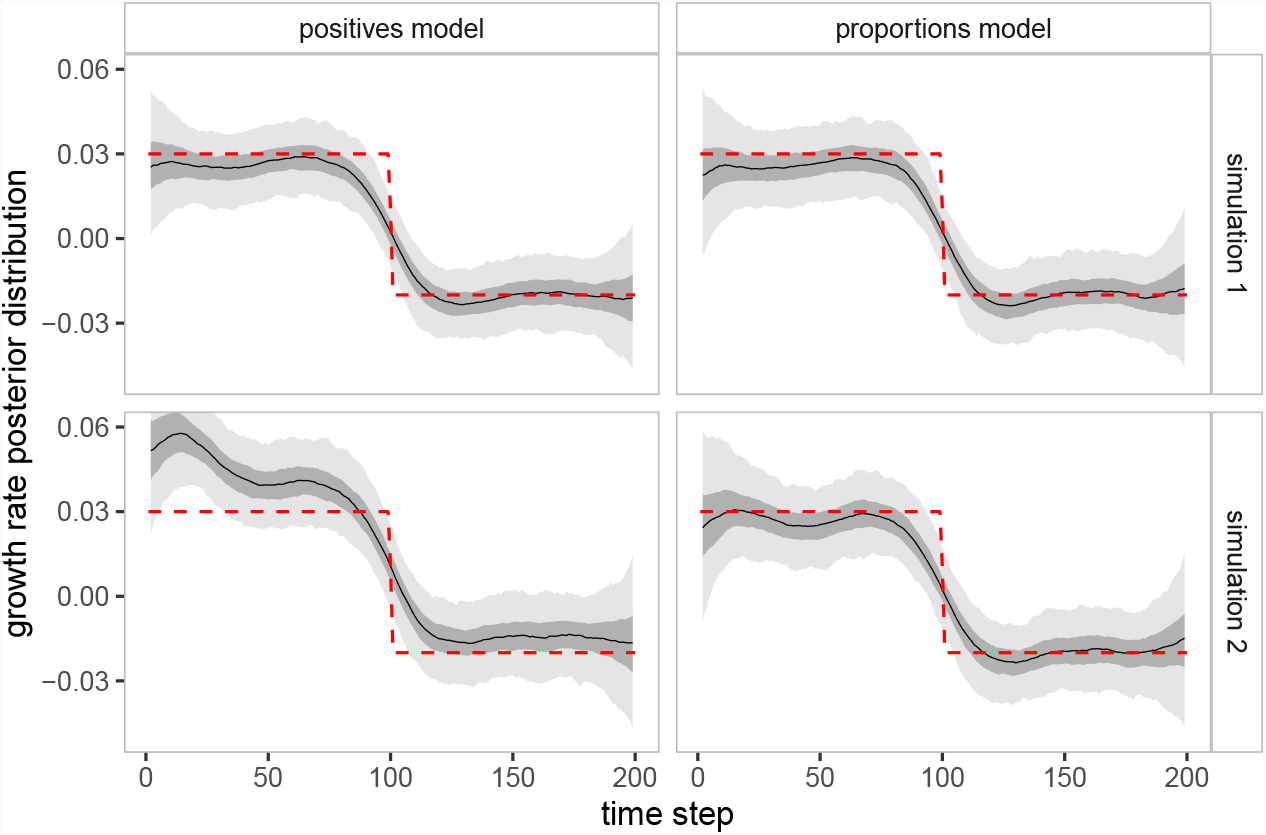
Validation of the ‘positives model’ and the ‘proportions model’. Posterior distributions of the growth rate for simulated data under two scenarios (top - constant testing; bottom - increasing testing) and two models (left - positives model; right - proportions model). We display the median (solid black line), 50% credible interval (dark shaded ribbon) and 90% credible interval (light shaded ribbon). The red dotted lines indicate the true growth rate.

**FIGURE 2.**
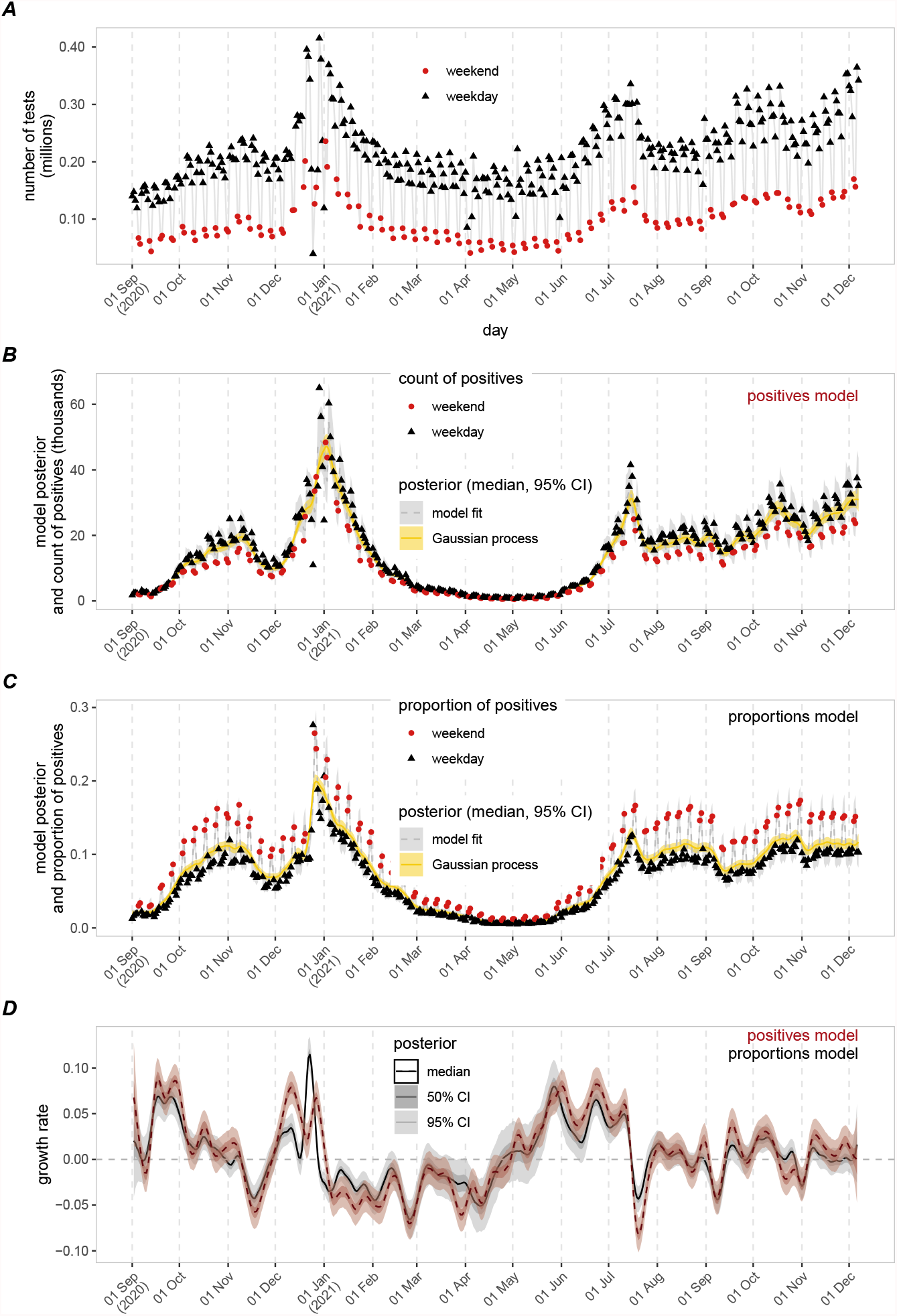
Model fitting and posterior distribution of the growth rate for SARS-CoV-2 cases in England from 1st September 2020 to 6th December 2021. (Panel A) Number of tests conducted. Black triangles correspond to reported test counts on weekdays, whilst red circles correspond to reported test counts on weekends. (Panel B) Median (lines) and credible interval (darker shaded ribbons for 50%, lighter shaded ribbons for 95%) of the model fitting (grey, dashed line) and the Gaussian process (yellow, solid line) for the positives model. Dots correspond to the daily count of positives. (Panel C) Median (lines) and credible interval (darker shaded ribbons for 50%, lighter shaded ribbons for 95%) of the model fitting (grey, dashed line) and the Gaussian process (yellow, solid line) for the proportions model. Dots correspond to the proportions of positives per day. (Panel D) Median (lines) and credible interval (darker shaded ribbons for 50%, lighter shaded ribbons for 95%) for the growth rate estimations in the positives model (red, dashed line) and proportions model (black, solid line).

For simulation 2, which has a linearly increasing daily testing rate, the positives model generally overestimates the growth rate. The overestimation in the positives model is more dramatic for the first 100 time steps, where the exponential growth rate of testing was higher (Figure 1, bottom left panel). In contrast, for the proportions model the true growth rate lies within the posterior credible interval for the majority of time steps, as in simulation 1 (Figure 1, bottom right panel).

### 2.5 Heterogeneity measure

Although the method is not inherently spatial, treating each set of temporal data as statistically independent, we can nevertheless use the spatial position of each spatially sampled location to address localised effects. In this way, we introduce a heterogeneity measure to assess whether exceptionally high or low growth rates within a given spatial location are a localised pattern or are caused by a larger, more widespread, phenomenon. We define the *heterogeneity h*_*i*_ of a spatial element *i* as:

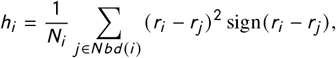

where *r*_*i*_ is the growth rate within location *i, j* ∈ *N bd* (*i*) denotes all spatial locations that neighbour element *i* (where for simplicity we assume this means share a boundary, but could be any measure of spatial locality) and *N*_*i*_ is the number of neighbours of *i*. Samples of *h*_*i*_ are taken by sampling from *r*_*i*_ and *r*_*j*_. As such, *h*_*i*_ provides a measure of local covariance, with its sign reflecting whether it has higher or lower growth than its neighbours. Moreover, we can estimate other quantities that allows us to compare the heterogeneity measure of different spatial elements. For instance, we estimate Prob(*h*_*i*_ > 0), allowing us to identify elements with considerably high heterogeneity.

## 3 CASE-STUDY

### 3.1 Data

We apply the models described above to data on daily counts of SARS-CoV-2 cases in England and in *lower tier local authorities* (LTLAs) between 1st September 2020 and 6th December 2021, dataset provided by Public Health England (now UKHSA). The data correspond to the count of people from the wider population (Pillar 2 of the UK government testing programme (UK Health Security Agency, 2021)) with at least one positive PCR test, reported by specimen date and residence location (by LTLA). The data also include the count of negative tests. The dataset includes a total of 4.51 million positives cases (timeseries shown in Figure 2, panel B), with test positivity ranging between 0-0.3 (Figure 2, panel C), and a total test count of 62.8 million (timeseries shown in Figure 2, panel A). We applied the models at a national-level, to case counts in England (Section 3.2), and at a local-level, to cases per LTLA in England (Section 3.3).

### 3.2 Growth rate estimation of SARS-CoV-2 in England from 1st September 2020 to 6th December 2021

We apply the positives and proportions model to the count of cases of SARS-CoV-2 in England between 1st September 2020 and 6th December 2021. For both models, we chose *l*_0_ = 100 days and *τ*_0_ = 1 as the baseline value for the length-scale and precision, respectively, and *B* = 𝕀. We replicate the same choices for *h* for the approximation of the derivative as in Section 2.4. Following the implementation details in Section 2.3, the model takes less than 3.0 seconds of CPU time to estimate the posterior distribution of the parameters (using the package `INLA` 21.07.10 in R 4.1.0 using a MacBook Pro with the Apple M1 chip) and an additional 4.7 seconds to generate 1000 samples of the parameters.

Our time period of study contains both the second wave of infections (punctuated by a short-term imposition of strong non-pharmaceutical interventions from 5th November to 2nd December 2021) and the protracted third wave. It is clear from the data that there is a pronounced effect of weekends on the testing patterns, with lower testing but a higher proportion of positives on a weekend (shown as red circles in Figure 2).

Both the fitted positives and proportions model had reasonable correspondence with the empirical data (Figure 2, panels B&C). The yellow ribbon shows the credible interval of the underlying Gaussian process, while the grey ribbon shows the model fit including day-of-the-week effects. Our posterior distributions for the hyperparameters of the Gaussian process were confined to a narrow region of the prior distribution, showing we had garnered knowledge from the available data (Figure 3, panel A). For the positives model, the standard deviation *σ* had a posterior median of 5.97 (95% credible interval 3.93-10.88) and the length scale *l* had a posterior median of 120.97 (95% credible interval 89.12-187.73). The proportions model had a similar pattern with lower values, where the standard deviation *σ* had a posterior median of 2.26 (95% credible interval 1.56-3.81) and the length scale *l* had a posterior median of 59.34 (95% credible interval 43.71-91.34).

**FIGURE 3.**
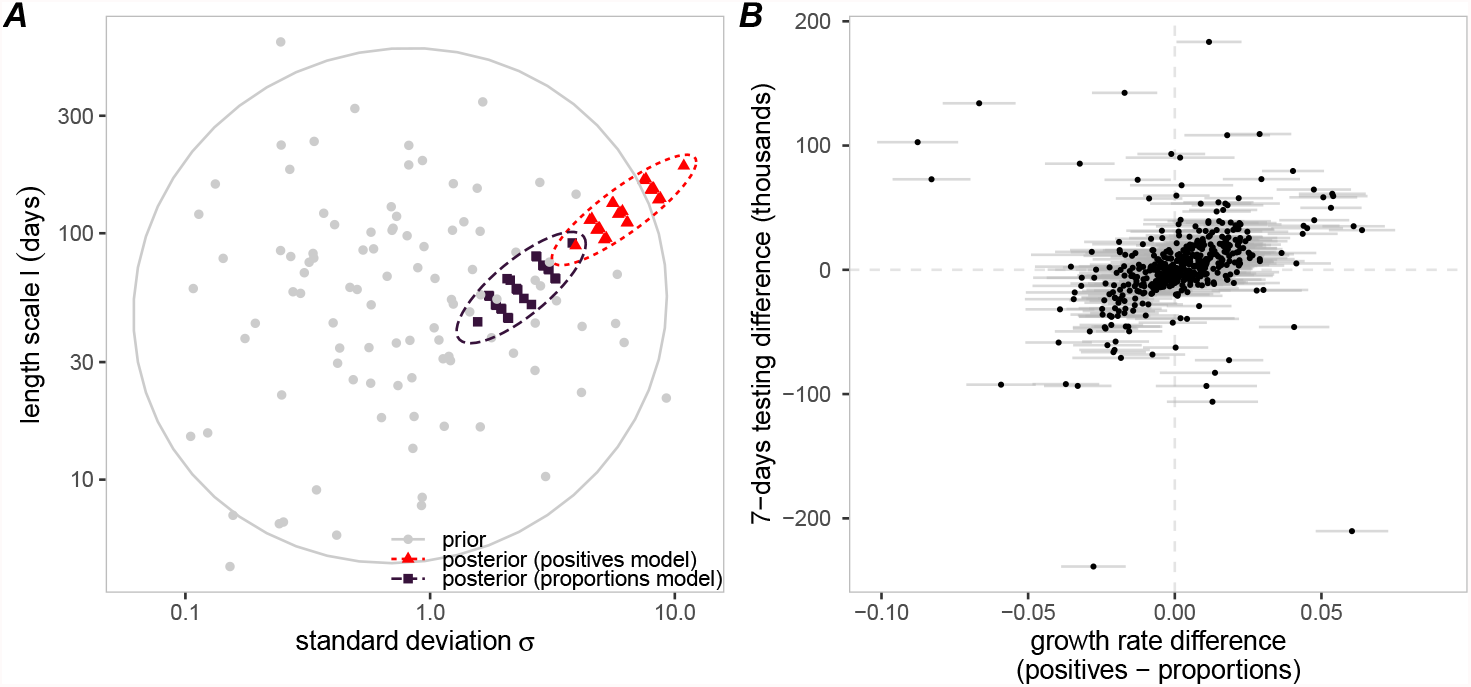
(Panel A) Comparison of the prior and posterior distributions of length-scale *l* (in days) and standard deviation *σ* for the positive and proportion models when applied to SARS-CoV-2 cases in England. Grey filled circles correspond to samples from the prior distribution. Red triangles correspond to samples from the posterior distribution in the positives model. Black squares correspond to samples from the posterior distribution in the proportions model. The dashed ovals represent the 95% posterior density region of each distribution. (Panel B) Comparison between the difference in testing (change in the count of tests in 7 days, y-axis) and the difference between the growth rate estimations of the positives and the proportions model (x-axis). The filled circle markers correspond to the median growth rate difference between the two models, with horizontal bars representing the 95% credible interval of the difference between growth rates.

There was usually a high level of concordance in the qualitative relationship between the growth rate estimates from the positives model and proportions model, with the models particularly well-agreeing whether the growth rate was positive or negative (Figure 2, panel D). This agreement provides additional confidence that we are seeing a robust signal from the data. Nevertheless, there were sustained periods with the two models producing dissimilar quantitative estimates, such as during December 2020. Higher differences in testing correspond to higher differences in growth rate estimation (Figure 3, panel B). This helps explain the discrepancies observed in December 2020, when testing practices are likely to be affected by the holiday period.

### 3.3 Spatial heterogeneity in cases of SARS-CoV-2 in the North-West region in England, April 2021

We applied the proportions model to the count of positive cases of SARS-CoV-2 in England for each of the 317 LTLAs. Since data at a lower resolution can be noisy, setting weak priors for the hyperparameters of the Gaussian process can lead to unrealistic length scales to account for the noise. To overcome that issue, we assume that the covariance function of the underlying Gaussian process at a local authority level has a similar shape to the national data. Therefore, we set the baseline values *σ*_0_ and *l*_0_ for the LTLA level to be the posterior median of *σ* and *l* obtained with the national data in Section 3.2, respectively, with precision *B* = 10𝕀.

We focus on the results from 23rd April 2021, when infections with the Delta variant were increasing in the North-West of England, particularly in Bolton where our proportions model gave an estimated positivity of 3.25% (95% PI: 2.65%-3.90%) (Figure 4, panel A). Multiple neighbouring LTLAs in the North-West region had median estimates for proportion of tests being positive above 2%. In other regions at that time, some urban centres had a similar high incidence of 2% or above, which included Manchester and Sheffield. However, we generally measured incidence to be lower in other regions compared to the North-West. For example, all LTLAs in the South-West and along the southern coast had low median incidence estimates (below 1%).

**FIGURE 4.**
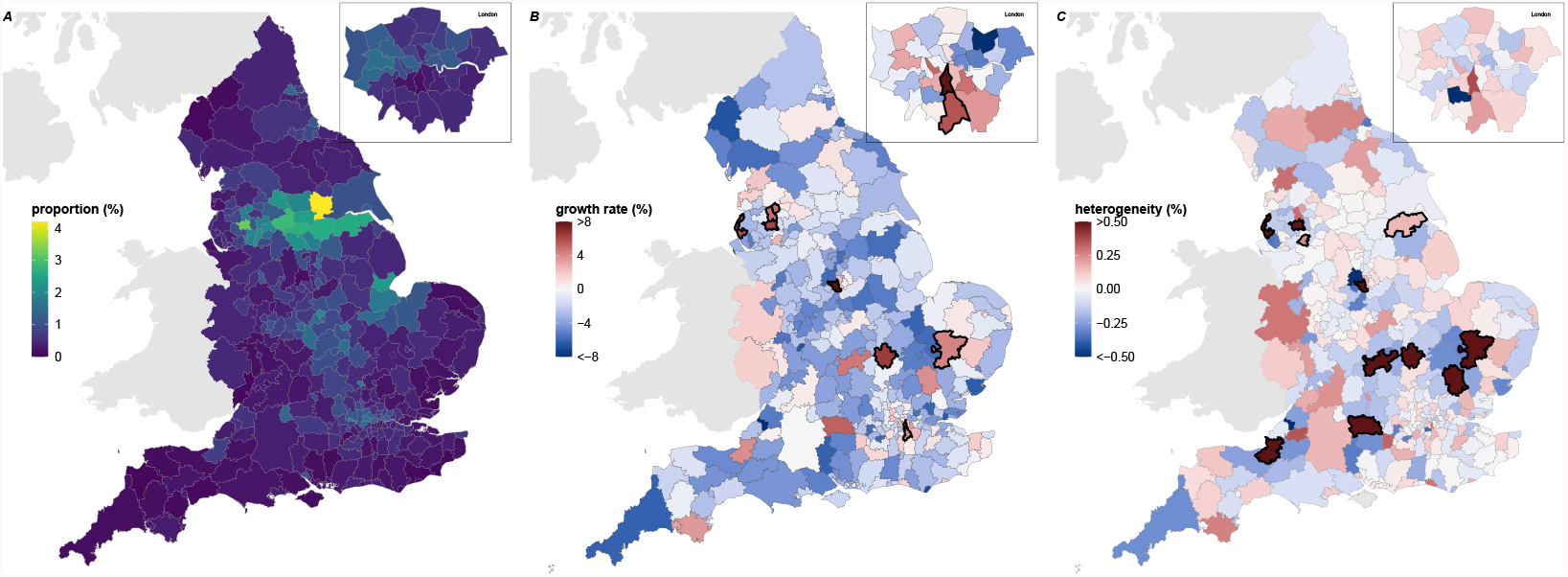
Epidemiological trends at the LTLA level in England on 23rd April 2021. (left) Estimated median of posterior of incidence, with lighter shading corresponding to higher incidence estimates. (center) Estimated median of posterior of growth rate (red shading when greater than zero, blue shading when less than zero). Regions with thicker borderlines correspond to LTLAs where the probability that the growth rate is greater than 0 exceeded 95%. (right) Median of heterogeneity (red shading when greater than zero, blue shading when less than zero). Regions with thicker borderlines correspond to LTLAs where the probability that the heterogeneity is greater than 0 exceeded 95%.

Though there was regional structure to the magnitude of test positivity, for growth rates we observed spatial variability in areas experiencing high growth in cases and those where incidence was declining (Figure 4, panel B). Areas expressing the greatest heterogeneity were regionally disconnected (Figure 4, panel C). LTLAs whose probability of positive heterogeneity exceeded 0.95, thereby indicating high growth rates larger than the surrounding areas, included Erewash in the East (median heterogeneity: 1.42), Sefton in the North-West (median heterogeneity: 1.00), Bedford in the east (median heterogeneity: 0.79), and Bolton in the North-West (median heterogeneity: 0.50). For Bolton, our heterogeneity measure suggested that area was having a localised increase (>99% probability of heterogeneity being greater than 0) rather than a regionally-driven event.

Through concurrently considering the growth rate and the proportion of tests with a positive result, we could discern those LTLAs suffering from both high prevalence and high growth rates (thereby possibly requiring further support), such as Bolton and Blackburn with Darwen, and LTLAs to monitor closely due to having low prevalence but high growth rates, including Erewash, Bedford and Sefton (Figure 5, panel A). Although Selby has the highest estimated proportion testing positive (4.19%), the growth rate had been decreasing in the prior week (Figure 5, panel B).

**FIGURE 5.**
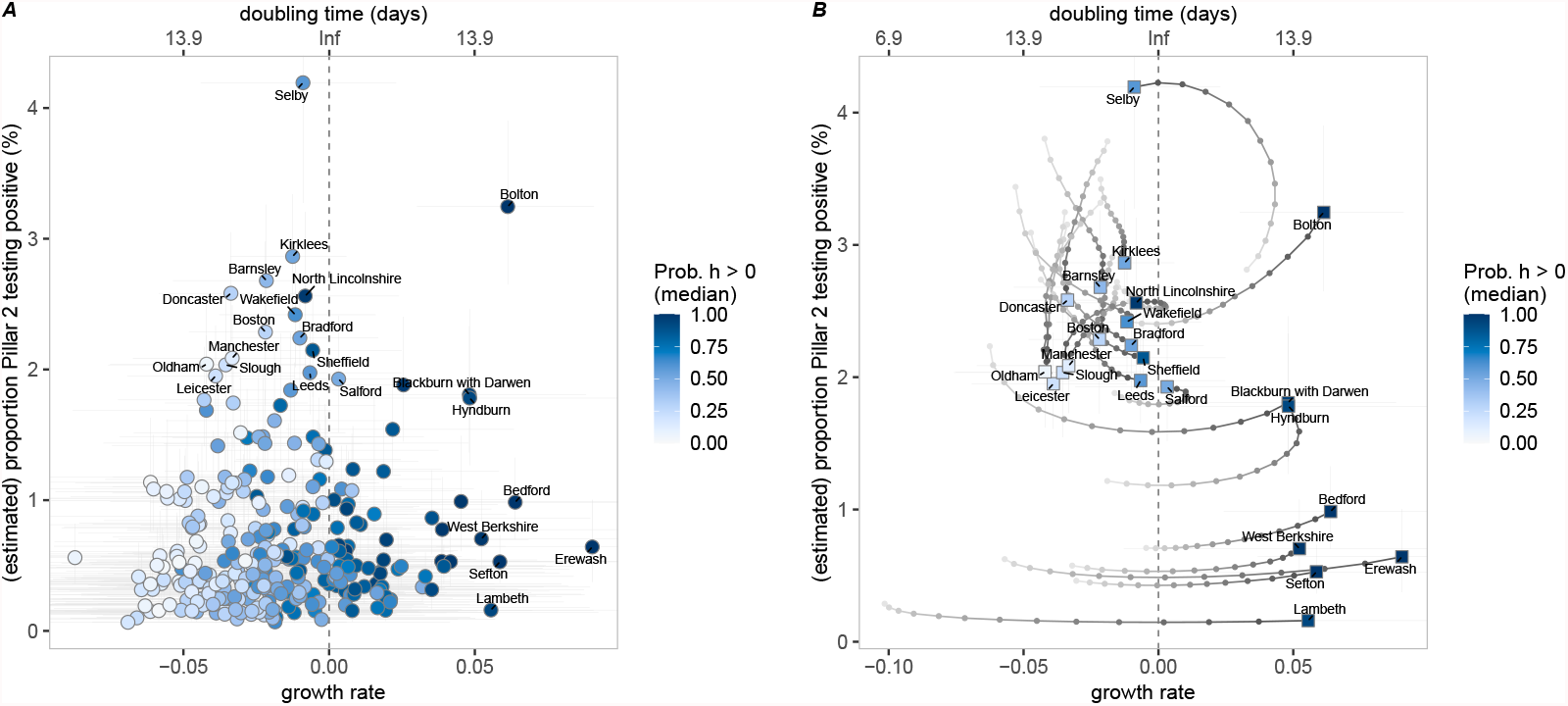
(left) Smooth estimation of positivity (y-axis) and growth rate (x-axis) of every LTLA in England on 23rd April 2021 coloured by probability that the heterogeneity is greater than 0 (dark blue for high probability, white for low probability). On the top axis we state the doubling time associated with the corresponding growth rate. Vertical bars correspond to the 95% credible interval of positivity. Horizontal bars correspond to the 95% credible interval of the estimated growth rates. (right) Trajectory of incidence-growth rate for LTLAs with high prevalence (top 2.5%) or high growth rate (top 5%) from 15th April 2021 and 23rd April 2021 (squares correspond to the 23rd April).

## 4 DISCUSSION

In this paper, we have proposed two model structures, the positives model (which only uses data on confirmed positive cases) and the proportions model (which uses both positive and negative test information), to estimate the instantaneous growth rate of cases. We note that any measure based on cases is necessarily a lagged indicator of infectious processes due to the delay between infection and notification of disease, which generally only occurs once symptoms arise. However, as we show for simple models, our methods can robustly estimate both the growth rate and temporal changes in the growth rate, which are often related to external epidemiological factors of public-health interest.

The latent structure of both models includes a Gaussian process (GP) that interpolates the epidemic curve and approximates the underlying process that generates the disease incidences. We then take samples of the derivative of the GP to estimate the growth rate. The models are implemented using the Laplace approximation incorporated in the `INLA` package in R. Both models use data on positive reported infections, while the proportions model also incorporates testing counts, enabling us to account for changes in test-seeking behaviour. We believe our approach has four benefits over existing methods. Firstly, it is rapid, robust and computationally efficient - all of which are considerable advantages when dealing with a rapidly changing epidemic in multiple spatial locations. Secondly, by focusing on growth rate rather than the reproduction number, we by-pass the complexities of estimating generation-time distributions that can substantially hinder other methods early in an outbreak. Thirdly, the combined use of positive reported infections and number of tests allows us to deal with the proportion of tests that are positive, a measure that is relatively insensitive to changes in testing behaviour. Finally, the use of Gaussian processes means that the method is also relatively robust to missing data, allowing us to provide continuous estimates even if some of the data streams are considered unreliable (for instance, the high rate of false negatives reported by the Immensa Health Clinic in some regions in the UK in September 2021 (Torjesen, 2021)).

Throughout we applied our method to reported cases of SARS-CoV-2 infection in England as confirmed by PCR testing. We perform our analysis both at a national scale (Figure 2) and at a small regional scale (Figures 4 and 5). Our choice of pathogen was determined by the need to quantify and explain the ongoing pandemic, feeding our findings through SPI-M-O (Scientific Pandemic Influenza Group on Modelling, Operational sub-group) to policy advisers. England has seen three major waves of infection, broadly associated with the wild-type, Alpha and Delta variants. The first wave which began in March 2020 led to large numbers of hospital admissions and deaths, but was poorly quantified in terms of infection due to the low level of community testing. The second wave began in September 2020 and peaked in late December 2020 or early January 2021 with over 60,000 cases reported on 29th December 2020. The third wave from June 2021 has been characterised by a prolonged period (over 5 months) of high cases, but with relatively low hospital admissions and deaths due to high vaccine uptake.

The national trends in growth rate highlight the complex pattern of growth (*r* > 0) or decay (*r* < 0) over time (Figure 2). Some notable changes that correspond to mitigation activities include: a pronounced negative growth rate during November 2020 due to the National 4-week lockdown, although the growth rate had been lower in October 2020 than September 2020; the negative growth rate during January-April 2021, during which time England was in lockdown followed by Steps 1 and 2 of the government’s COVID-19 response (UK Cabinet Office, 2021), which transitioned into high growth rates by late May 2021; a sharp drop in growth rate (especially as estimated by the positives model) in July 2021 which has been labelled as the ‘pingdemic’ due to the large number of individuals contacted through the Test-and-Trace App, and the potential changes in behaviour to avoid this; finally, we observe that much of August-November 2021 is characterised by growth rates close to zero, reflecting the high level of cases that have been maintained through this period.

Both the positives model and proportions model aim to capture the instantaneous growth rate of new cases and, if the efforts in testing are constant, both methods provide equivalent results. However, the estimations can differ when testing behaviour has a temporal trend - as seen during the COVID-19 outbreak in England. For instance, if the testing rate increases, the positives model can underestimate the actual growth rate (Figure 3, panel B). In contrast, the proportions model accounts for changes in the number of tests and can give more reliable estimates. However, both models can be affected by more nuanced changes in testing behaviour; our proportions model assumes that any change in test-seeking behaviour affects all sections of the population equally - if this is not true (such as the introduction of twice-weekly lateral flow testing for secondary school children) then there can be biases. We propose to include both approaches into routine analysis since they give different perspectives to the same data, particularly when there is little knowledge of the processes driving testing behaviour in the population.

Another strength of our growth rate estimation method is the relatively low computational expense and run time, using the Laplace approximation implemented in `INLA` (Rue et al., 2009), permitting the application of the model at a local level (to each of the 317 LTLAs in England). Spatially, the English COVID-19 case data is either broken into seven National Health Service regions, or into 317 Lower Tier Local Authorities (LTLA). LTLAs range in size from just over 2000 people (Isles of Scilly) to well over a million (Birmingham), but most contain around 140,000 inhabitants. Performing our analysis at this spatio-temporal scale allows us to identify both highly localised outbreaks (as seen in the maps in (Figure 4) or wider regional trends, enabling scrutiny of locations exhibiting atypical data patterns. Furthermore, introducing a heterogeneity measure enabled comparisons of the growth rates between neighbouring LTLAs. The heterogeneity measure has been used during the pandemic to highlight places with abnormal growth patterns, generally identifying LTLAs with significantly higher growth. The process has also be extended (by considering S-gene target failure) to quantify the spread of new variants (e.g. Alpha and Delta) to pinpoint localities that were increasing above mere noise (Challen et al., 2021).

Our analysis of LTLAs was focused around 23rd April 2021; at this time the Delta variant had begun to establish across England (with about 20% of cases attributed to Delta), hospital admissions and deaths were continuing to decline, but community cases had reached a nadir. Understanding the spatial patterns of growth at this time, and linking it to the prevalence of the Delta variant, was important for assessing the invasion of the new variant. We observe a mixed mosaic of growth rates across England (Figure 4) with a few regions where the growth rate is significantly above zero. Many of these regions also appear in the heterogeneity map as islands of growth amid a sea of declining cases; which suggests a rapid localised growth in these areas. Focusing on LTLAs that either have high growth rates or high prevalence (Figure 5) we identify three main grouping that may require further epidemiological investigation. Firstly there are four LTLAs (South Hams, South Northamptonshire, Erewash and Hyndburn) that have high positive growth rates and where we expect cases to continue to rise. Secondly, there is a group of fifteen LTLAs where a high proportion of tests (between 2-4%) are positive; of these Bolton, Trafford and again Hyndburn (all in the North-West of England) are of the greatest concern due to their positive growth rate. Finally, Selby in the North-East of England (clearly identifiable on the incidence map of Figure 4) has an extremely high proportion of tests that are positive, and while the mean growth rate is slightly below zero this is not statistically significant suggesting that cases will remain high over the short-term.

Our approach for estimating the growth rate is a purely statistical method and therefore has limitations. First, the model is non-mechanistic and does not incorporate any epidemiological assumptions. Therefore, it is not suitable for predicting future changes in infections or making long term forecasts, particularly as it cannot account for the depletion of susceptible through infection or vaccination. Second, we assume that the spatial regions investigated are independent and homogeneous, we do not account for the movement of infection between regions (Kraemer et al., 2021) nor the spatial and social structure within a region. A lack of internal structure could be important for public-health concerns; for example, an outbreak that is primarily increasing in the young has very different health implications compared to one that is increasing in the elderly. There is no reason why richer data structures cannot be incorporated within our methodology (for example looking at the growth rate in a set of age-groups), but such an analysis requires large amounts of data and is increasing complex to interpret. Third, the data analysed in this study comes from PCR testing (or individuals that have performed a lateral flow test followed by PCR). Therefore, there are limitations due to specificity and sensitivity of the test and the ability of individuals to swab reliably. Associated with this, and discussed above, changes to test-seeking behaviour beyond a simple increase in testing could introduce a range of biases. It is important to stress that throughout we are fitting to positive tests not infections, although we believe the two are highly correlated. Finally, though Gaussian processes provide a flexible tool, some prior knowledge of the patterns of the disease is required to inform the subjective choice of the covariance function and its priors. If the data sources are not consistent over the time course of the study, it will affect both models. Moreover, abrupt changes in the epidemic curve are harder to pick for certain covariance functions (e.g. smooth covariance functions). This highlights the need for further studies around how to design more complex covariance functions that allow such abrupt changes to be captured.

In summary, we have presented a general structure for estimating instantaneous growth rates that uses a Bayesian hierarchical model to fit a Gaussian process to the epidemiological data. Applied to high-resolution spatio-temporal SARS-CoV-2 case and testing data from England, we have demonstrated the ability of parsimonious models estimating instantaneous growth rate to both determine long-term patterns of growth at a national-scale, and highlight growth and spatial heterogeneity at a regional-scale.

## Data Availability

Data is available to researchers with data sharing agreements in place with Public Health England.

## ACKNOWLEDGEMENTS

We are grateful to Nick Gent and Public Health England (now the UK Health Security Agency) for providing access to data on positive and negative PCR tests by LTLA. The ethics of the use of these data for these purposes was agreed by the UK Health Security Agency with the Government’s SPI-M(O) / SAGE committees.

## CODE AVAILABILITY

Code for the study is available at GitHub/juniper-consortium/growth-rate-estim.

