## Supplementary material for "Bayesian Estimation of real-time Epidemic Growth Rates using Gaussian Processes: local dynamics of SARS-CoV-2 in England"

### S | SUPPLEMENTARY MATERIAL

#### S.1 | Directed acyclic graph of the models

We display directed acyclic graph corresponding to the positives model described in Section 2.1.1 (Figure 1) and the proportions model described in Section 2.1.2 of the main document (Figure 2), respectively.

#### S.2 | Growth rate comparison

Let $w$, $n$ and $z$ be continuous functions on $\mathbb{R} \cup \{0\}$, such that at a given time $t \in \{0, 1, \dots\}$, $w(t)$ denotes the number of new cases, $n(t)$ denotes the number of tests, and $z(t)$ denotes the number of positive tests. Note that although $w$, $n$ and $z$ are continuous functions, their values have an interpretable meaning only on discrete times (for instance, daily counts). Our goal is to estimate the growth rate $r(t)$, defined as the *per capita* change in the number of new cases per time; that is, $r(t) = \partial_t(w(t))/w(t)$.

In the positives model, we approximate $r(t)$ as the growth rate of observed positive tests $z(t)$, denoted $r_z(t) = \partial_t(z(t))/z(t)$. We describe $z(t)$ in terms of a latent function $x(t)$ such that $z(t) = \exp(x(t))$, which simplifies the growth rate as $r(t) \approx r_z(t) = \partial_t(x(t))$.

In the proportions model, we describe the proportion of positive tests $z(t)/n(t)$ in terms of a latent function $x(t)$ such that $z(t)/n(t) = \text{logit}^{-1}(x(t))$. The derivative of $x_t$ is not directly related to $r_z(t)$ as in the positives model; however, we show below it is related to $r_z(t)$ and $r_n(t)$, where $r_n(t) = \partial_t(n(t))/n(t)$ is the growth rate of number of tests performed. First, we

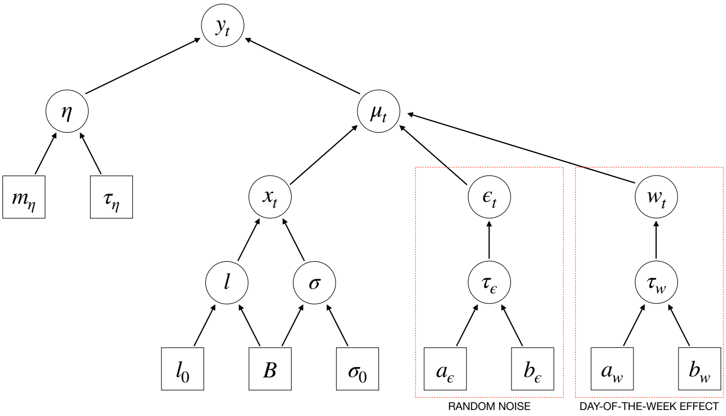


FIGURE 1 Directed Acyclic Graph describing the hierarchical conditional independence structure of the positives model, described in Section 2.1.1. The parameters $\eta$, $\mu_t$, $x_t$ and $\epsilon_t$ and the hyperparameters $l$, $\sigma$, $\tau_\epsilon$ and $\epsilon_w$ are enclosed in circles. Inputs of the model are enclosed in squares.

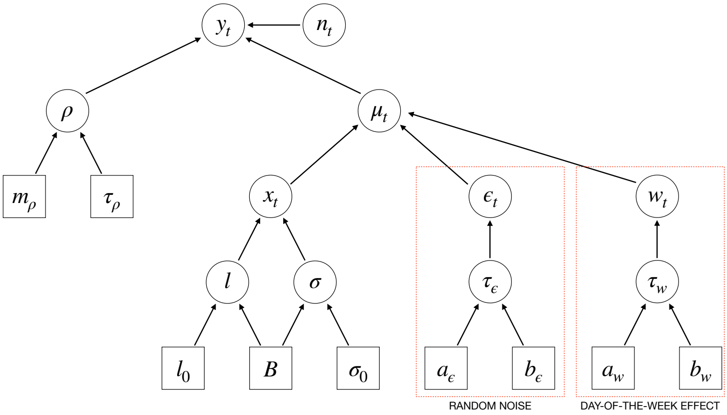


FIGURE 2 Directed Acyclic Graph describing the hierarchical conditional independence structure of the proportions model, described in Section 2.1.2. The parameters $\rho$, $\mu_t$, $x_t$, $\epsilon_t$ and $w_t$ and the hyperparameters $l$, $\sigma$, $\tau_\epsilon$, $\epsilon_t$ and $w_t$ are enclosed in circles. Inputs of the model are enclosed in squares.

compute the derivative of the function $x(t)$:

$$
\begin{aligned}
\partial_t(x(t)) &= \partial_t\left[\log\left\{\frac{z(t)}{n(t)-z(t)}\right\}\right] \\
&= \left(\frac{n(t)-z(t)}{z(t)}\right)\left(\frac{n(t)\partial_t(z(t)) - z(t)\partial_t(n(t))}{(n(t)-z(t))^2}\right) \\
&= \{r_z(t) - r_n(t)\}[1+\exp\{x(t)\}].
\end{aligned}
$$

Then, we approximate $r(t)$ as the growth rate of positive tests minus the growth rate of number of tests: $r(t) \approx r_z(t) - r_n(t) = \partial_t(x_t)/[1+\exp\{x(t)\}]$.
